# Covid-19 vaccination decisions and impacts of vaccine mandates: A cross sectional survey of healthcare workers in Ontario, Canada

**DOI:** 10.1101/2024.06.23.24309372

**Authors:** Claudia Chaufan, Natalie Hemsing, Rachael Moncrieffe

## Abstract

**Background:** Since vaccination policies were introduced in the healthcare sector in the province of Ontario, Canada, most establishments implemented vaccination or termination requirements, with most enforcing them to this day. Researchers have shown a strong interest in the perceived problem of vaccine hesitancy among healthcare workers, yet not in their lived experience of the policy or in their views on the policy’s impact on the quality of patient care in the province.

**Goal:** To document the experience and views on mandated vaccination of healthcare workers in the province of Ontario, Canada.

**Methods:** Between February and March 2024, we conducted a cross-sectional survey of Ontario healthcare workers, recruited through professional contacts, social media, and word-of-mouth.

**Findings:** Most respondents, most with 16 or more years of professional experience, were unvaccinated, and most had been terminated due to non-compliance with mandates. As well, and regardless of vaccination status, most respondents reported safety concerns with vaccination, yet did not request an exemption due to their experience of high rejection rates by employers. Nevertheless, most unvaccinated workers reported satisfaction with their vaccination choices, although they also reported significant, negative impacts of the policy on their finances, their mental health, their social and personal relationships, and to a lesser degree, their physical health. In contrast, most respondents within the minority of vaccinated respondents reported being dissatisfied with their vaccination decisions, as well as having experienced mild to serious post vaccine adverse events, with about one-quarter within this group reporting having been coerced into taking further doses, under threat of termination, despite these events. Further, a large minority of respondents reported having witnessed underreporting or dismissal by hospital management of adverse events post vaccination among patients, worse treatment of unvaccinated patients, and concerning changes in practice protocols. Close to half also reported their intention to leave the healthcare industry.

**Discussion:** Our findings indicate that in Ontario, Canada, mandated vaccination in the health sector had an overall negative impact on the well-being of the healthcare labour force, on patient care, on the sustainability of the health system, and on ethical medical practice. Our study should be reproduced in other provinces, as well as in other countries that adopted comparable policies. Findings from this and similar studies should be seriously considered when planning for future health emergencies, to protect health systems in crisis due to severe labour shortages, as well as the right to informed consent of healthcare workers and members of the public.

## Introduction

When Covid-19 vaccination policies were introduced in the healthcare sector in the province of Ontario, Canada, tensions regarding mandates for healthcare workers (hereafter HCWs) became apparent, especially conflicting opinions regarding their impact on the labour force and subsequently patient care. Based on the guidance from the National Advisory Committee on Immunization (NACI), HCWs, along with other groups designated as high risk, were prioritized for vaccination during “phase 1” – December 2020 to March 2021 – of the vaccine rollout in the province (Fitzpatrick et al., 2022; Ismail et al., 2020). On August 17, 2021, the Ontario Chief Medical Officer of Health issued Directive #6, a policy that required hospitals and home and community care establishments to implement a Covid-19 vaccination policy for employees, contractors, students, and volunteers. The directive went into effect on September 7, 2021, and remained in effect through March 14, 2022, establishing a mandatory vaccination policy that allowed exemptions to individuals with an officially approved medical waiver or willing to take a test to learn about the ostensible benefits of vaccination before declining (Office of the Auditor General of Ontario, 2022). Nevertheless, most healthcare settings chose to implement vaccination as a condition of employment, with very limited medical exemptions, and no test-taking alternative, and the Ontario Ministry of Long-term Care issued a vaccine mandate effective December 2021, also through March 2022, that required all staff, contractors, students, and volunteers, to either be vaccinated or terminated.

While most health professional organizations and regulatory colleges also offered their broad support for vaccination policies, endorsement of *mandatory* vaccination for HCWs was mixed, with some stakeholders warning about potential negative impacts. Some of the tensions over the introduction of vaccination mandates for HCWs were reported in Canadian media sources (CBC News, 2022; McKenzie-Sutter, 2021). For example, when in October 2021, Ontario Premier Doug Ford asked for input from health leaders across the province, the Council of Ontario Medical Officers of Health, representing public health officials from the province’s 34 public health units, communicated their approval of a mandatory vaccination policy (McKenzie-Sutter, 2021). So did the Ontario Hospital Association, arguing that the policy would increase vaccine uptake, thus making healthcare settings safer (ibid). Supporters also included the Registered Nurses Association of Ontario (RNAO) that, in an open letter to the Premier, recommended “a clear, comprehensive [and] firm mandate”, because “patients, already vulnerable, [would] have a greater sense of comfort and safety knowing that they [would] not get COVID from [an unvaccinated] HCW”, and because “mandatory vaccination [was] considered by virtually all nurses […] necessary for the protection of their health and safety and those they go home to every day” (Registered Nurses Association of Ontario, 2021). RNAO CEO Doris Grinspun, who penned the letter, further proffered that “the absence of mandatory vaccinations is influencing decisions of nurses to leave workplaces and even the profession” (ibid).

From an opposing perspective, the Ontario Nurses Association (ONA) noted that introducing a policy that provided only the option to be vaccinated or terminated, when many nurses were already at their “breaking point”, would “exacerbate an already tenuous situation on the cusp of a catastrophic staffing crisis”, potentially reducing patient safety (McKenzie-Sutter, 2021). On a similar vein, as revealed by a FOI request the following year, the Hospital Notre-Dame CEO cautioned that a vaccine mandate could lead to reduced services, noting that hospital staff were “very well informed … and still choose not to be vaccinated”, and concluding that it would be best to “leave it to the individual hospital to decide” whether or not to mandate vaccination (CBC News, 2022). Finally, unions such as SEIU Healthcare, representing over 60,000 frontline HCWs, urged the Premier to focus on other major factors, such as low wages, to address an impending staffing crisis, with SEIU Healthcare president, Sharleen Stewart noting, in a joint statement with the Ontario Council of Hospital Unions, that “more [HCWs] are leaving the system because of poor wages and working conditions” rather than because of a concern with insufficient vaccination among their co-workers (McKenzie-Sutter, 2021).

Be that as it may, vaccine mandates for HCWs have since received broad support and have been enforced by most medical establishments, with many enforcing them to this day. They have also been supported by research addressing the perceived problem of “vaccine hesitancy” by, for instance, identifying HCWs’ personal characteristics – psychological, cultural, ideological - that may explain their less than full embrace of vaccination (Achat et al., 2022; Evans et al., 2022; Lee et al., 2022; Oberleitner et al., 2022; Ulrich et al., 2022), and subsequently recommending interventions such as “targeted messaging” (Oberleitner et al., 2022), better information or education (Dietrich et al., 2022) or, as a “last resort”, mandatory vaccination (Lee et al., 2022). This line of research has also framed those who doubt or resist the policy as uneducated or misinformed (Achat et al., 2022; Dietrich et al., 2022), and in all cases troublesome, especially given their role as trusted sources of vaccine information (Evans et al., 2022). As a result, there has been limited research on the vaccination experiences or perspectives on mandates of HCWs themselves, free from preconceptions about the desirability of vaccination (Chaufan & Hemsing, Forthcoming). To address this gap, we surveyed HCWs in Ontario, Canada, documenting their experience of workplace vaccination mandates and their views about the impact of the policy on healthcare system sustainability and quality.

## Methods

We conducted an online survey of Ontario HCWs. Eligibility criteria included anyone working in a healthcare setting – whether in patient care, administrative, or maintenance services – in Ontario, and of any age and vaccination status. We advertised the study via the professional networks and contacts of the lead author. In turn, these contacts advertised it through social media. We also used a snowball sampling method whereby respondents were invited to disseminate recruitment materials among their own networks. Materials were resent at seven-day intervals over one month (McDonald et al., 2024). Based on the 918,700 people estimated to be employed in Ontario in the healthcare and social assistance industry in 2021(Government of Canada, 2024), we sought to recruit a convenience sample of 400 HCWs, for a confidence level of 95% and a margin of error of 5% (SurveyMonkey, n.d.). We exceeded our goal and included 468 HCWs.

We pilot tested the survey with a sample of 5 HCWs and integrated their feedback prior to launching it in February 2024, collecting responses through March 2024. The survey consisted of 15 sections, with sections automatically skipped depending on vaccination status or job termination for non-compliance with mandatory vaccination. After informing respondents about the purpose of the research and the confidentiality of the data, confirming that they worked in Ontario, and obtaining informed consent, they were asked about their employment status and history, their experience of making vaccination decisions, the impact of the policy on their finances, personal and social relations, and mental and physical health, and their perspectives on the impact of the policy on patient care. The survey consisted of 88 questions, including multiple choice, short answer, and Likert scale (i.e., rank ordered responses). Sections included: demographics (8 questions); employment (5 questions); Covid-19 experiences (2 questions); informed consent (11 questions); vaccination decision making (3 questions); vaccine side effects (3 questions); accommodations (3 questions); personal impact of vaccination policies (9 questions); self -rated health changes (4 questions); vaccination requirements and employment status (2 questions); impacts of job termination (10 questions); impacts on patient care (23 questions); experiences of administering Covid-19 vaccines (4 questions); and an open ended question for further comments (1 optional question). Following the survey, respondents were entered into a raffle for a $100 gift card. Due to the exploratory nature of the research, we performed only a descriptive analysis of the data using Excel spreadsheets. The research team met regularly to review the data, discuss the analysis, and identify trends. The study was approved by the York University Office of Research Ethics.

## Findings ^1^

### Demographics

Most respondents (273/468, 58%) were between the ages of 35-44 (138/468, 29.5%) and 45-54 (135/468, 29%), followed by ages 55-64 (105/468, 22.4%), ages 25-34 (54/ 468, 12%), and over 65 years of age (15/468, 3.2%). Most were women (385/468, 82.3%), a minority were men (52/468, 11.1%), a small minority (2/468, 0.4%) identified as another gender (transgender woman; non-binary), and the rest did not report their gender (9/468, 2%). Most respondents (286/468, 61.1%) self-identified as middle income, followed by roughly the same proportion of low (64/468, 14%) and high middle (59/468, 13%) income, and a small proportion of very low (14/468, 3%) and very high (7/468, 1.5%) income. Most reported Canada as their country of birth (360/468, 77%), followed by Poland (9/468, 2%), the USA (5/468, 1.1%), and Romania (5/468, 1.1%). Most respondents were Caucasian / White (392/ 468, 84%), followed by Latin American (14/468, 3%), Black (14/468, 3%), Indigenous (11/468, 2.4%), South Asian (9/468, 2%), and Chinese (5/468, 1.1%). Most (338/468, 72.2%) were married or living with a partner and about one-fifth (91/468, 19.4%) were either single (82/468,18%) or widowed (9/468, 2%). Most respondents (349/468, 74.5%) reported caretaking responsibilities for children or stepchildren (264/468, 56.4%) or parents (85/468, 18.2%), while close to one-third (139/468, 30%) reported no caretaking responsibilities (Table 1).

**Table 1.** Demographic characteristics^1^.

| **Category** | **Value** | **N** | **%** |
| --- | --- | --- | --- |
| **Age** | 18-24 | 0/468 | 0% |
|  | 25 to 34 | 54/468 | 12% |
|  | 35 to 44 | 138/468 | 29.5% |
|  | 45 to 54 | 135/468 | 29% |
|  | 55 to 64 | 105/468 | 22.4% |
|  | 65 or older | 15/468 | 3.2% |
|  | Total respondents | 447/468 | 96% |
|  | No response | 21/468 | 4.5% |
| **Gender** | Woman | 385/468 | 82.3% |
|  | Man | 52/468 | 11.1% |
|  | Prefer not to answer | 9/468 | 2% |
|  | Other (transgender woman; non-binary) | 2/468 | 0.4% |
|  | Total respondents | 448/468 | 96% |
|  | No response | 20/468 | 4.3% |
| **Country of birth** | Canada | 360/468 | 77% |
|  | USA | 5/468 | 1.1% |
|  | Poland | 9/468 | 2% |
|  | Romania | 5/468 | 1.1% |
|  | Other | 68/468 | 15% |
|  | Total respondents | 447/468 | 96% |
|  | No response | 21/468 | 4.5% |
| **Ethnic or Cultural Background** | Caucasian or White | 392/468 | 84% |
|  | Latin American | 14/468 | 3% |

<sup>1</sup> Some answers allow for multiple options, so totals do not always amount to 100%

|  |  |  |  |
| --- | --- | --- | --- |
|  | Black | 14/468 | 3% |
|  | Indigenous | 11/468 | 2.4% |
|  | South Asian (Indian, Pakistani, Sri Lankan, etc.) | 9/468 | 2% |
|  | Chinese | 5/468 | 1.1% |
|  | Other | 15/468 | 3.2% |
|  | No response | 22/468 | 5% |
| **Domestic status** | Married or living with a partner | 338/468 | 72.2% |
|  | Single | 82/468 | 18% |
|  | Widow (er) | 9/468 | 2% |
|  | Other | 18/468 | 4% |
|  | Total respondents | 447/468 | 96% |
|  | No response | 21/468 | 4.5% |
| **Caretaking responsibilities** | Children / Stepchildren | 264/468 | 56.4% |
|  | Parents | 85/468 | 18.2% |
|  | None | 139/468 | 30% |
|  | Other | 19/468 | 4.1% |
|  | No response | 18/367 | 4% |
| **Education level** | Bachelor of Science in Nursing (BScN) | 111/468 | 24% |
|  | Registered Nurse (RN) or Licensed Practical Nurse (LPN) or Registered Practical Nurse (RPN) diploma | 73/468 | 16% |
|  | Nursing Assistant Diploma | 9/468 | 2% |
|  | Master of Science in Nursing (MScN) or Master of Nursing (MN) | 13/468 | 3% |
|  | Nurse Practitioner (NP) graduate diploma | 4/468 | 1% |
|  | Doctor of Medicine (MD) | 12/468 | 3% |
|  | Paramedical or Emergency Medical Technician (EMT) program | 10/468 | 2.1% |
|  | Bachelor of Science in Midwifery | 5/468 | 1.1% |
|  | Bachelor of Health Science in Midwifery | 2/468 | 0.4% |
|  | Master of Science in Midwifery | 1/468 | 0.2% |
|  | Other Midwifery degree | 2/468 | 0.4% |
|  | Bachelor of Health Sciences (BHSc) | 12 /468 | 3% |
|  | Master of Health Sciences (MHSc) | 2/468 | 0.4% |
|  | Master of Public Health (MPH) | 2/468 | 0.4% |
|  | PhD (any field) | 7/468 | 1.5% |
|  | Bachelor of Health Science in Occupational Therapy | 1/468 | 0.2% |
|  | Master of Science in Occupational Therapy (MScOT) | 9/468 | 2% |
|  | Master of Health Sciences (MHSc) | 2/468 | 0.4% |
|  | Bachelor of Science in Physical Therapy (BScPT) | 1/468 | 0.2% |
|  | Master of Science in Physical Therapy (MScPT) | 1/468 | 0.2% |
|  | Bachelor or advanced degree, Health Administration/ Systems Management (e.g., BHAD) | 6/468 | 1.3% |
|  | Bachelor or Doctor of Pharmacy | 3/468 | 1% |
|  | Registered Pharmacy Technician (RPhT) | 7/468 | 1.5% |
|  | Bachelor of Science in Nutrition or Food Sciences/ Dietetics | 3/468 | 1% |
|  | Medical Radiation Technologist Diploma | 8/468 | 2% |
|  | Doctor of Dental Medicine/<br>Surgery (DDM, DDS) | 1/468 | 0.2% |
|  | Dental Hygiene diploma | 2/468 | 0.4% |
|  | Doctor of Optometry (OD) | 2/468 | 0.4% |
|  | Traditional Chinese Medicine<br>(TCM) diploma | 1/468 | 0.2% |
|  | Acupuncture diploma | 5/468 | 1.1% |
|  | Phytotherapy (herbal<br>medicine) diploma | 1/468 | 0.2% |
|  | Homeopathy | 2/468 | 0.4% |
|  | Other | 138/468 | 29.5% |
|  | No response | 20/468 | 4.3% |
| **Socioeconomic**<br>**status** | Very low-income | 14/468 | 3% |
|  | Low-income | 64/468 | 14% |
|  | Middle-income | 286/468 | 61.1% |
|  | High middle-income | 59/468 | 13% |
|  | High-income | 7/468 | 1.5% |
|  | Prefer not to answer | 20/468 | 4.3% |
|  | Total respondents | 450/468 | 96.2% |
|  | No Response | 18/468 | 4% |
| **Employment**<br>**status** | Employed full-time | 147/468 | 31.5% |
|  | Employed part-time | 97/468 | 21% |
|  | Unemployed | 70/468 | 15% |
|  | Self-employed | 70/468 | 15% |
|  | Casual | 16/468 | 3.4% |
|  | Contractor | 11/468 | 2.4% |
|  | Other | 37/468 | 8% |
|  | Total respondents | 448/468 | 96% |
|  | No response | 20/468 | 4.3% |
| **Ontario health region affiliation<sup>2</sup>** | Northern Region | 40/468 | 9% |
|  | Eastern Region | 85/468 | 18.2% |
|  | Southwest Region | 121/468 | 23% |
|  | Central Region | 76/468 | 16.2% |
|  | Toronto Region | 117/468 | 25% |
|  | Other | 13/468 | 3% |
|  | No response | 22/468 | 5% |
| **Profession/area of occupation** | Registered nurse/ Registered psychiatric nurse | 125/468 | 27% |
|  | Licensed practical nurse | 40/468 | 9% |
|  | Nurse aide/ Orderly/ Patient services associate (e.g., health care aide, long-term care aide, nursing assistant) | 28/468 | 6% |
|  | Nursing Coordinator/ Supervisor | 11/468 | 2.4% |
|  | Personal Support Worker (PSW) | 12/468 | 3% |
|  | Social Worker | 7/468 | 1.5% |
|  | Academic appointment/ University affiliation | 12/468 | 3% |
|  | In clinical practice | 74/468 | 16% |
|  | Not in clinical practice | 24/468 | 5.1% |
|  | Health profession student | 4/468 | 1% |
|  | Health professional in training (e.g., resident) | 1/468 | 0.2% |
|  | Specialist physician | 8/468 | 2% |

<sup>2</sup> Construction Forecasts. (n.d.). *Definitions of Ontario regions*. Definitions of Ontario Regions. Retrieved April 7, 2024, from https://www.constructionforecasts.ca/en/reference/definitions-ontario-regions#:~:text=The%20Southwest%20region%20includes%20the,Stratford%2C%20Goderich%20and%20Owen%20Sound.

|  |  |  |  |
| --- | --- | --- | --- |
|  | General practitioner/ Family physician | 5/468 | 1.1% |
|  | Allied primary health practitioner (e.g., nurse practitioner, midwife, physician assistant) | 10/468 | 2.1% |
|  | Paramedical occupation (e.g., EMT, ambulance attendant, advanced care paramedic) | 11/468 | 2.3% |
|  | Physiotherapist or Physiotherapy Assistant (PTA) | 4/468 | 1% |
|  | Occupational therapist or Occupational therapy Assistant (OTA) | 12/468 | 3% |
|  | Medical technologist/ Technician (e.g., medical laboratory technologist, respiratory therapist) | 32/468 | 7% |
|  | Dentist | 1/468 | 0.2% |
|  | Dental care - technical occupation (e.g., dental hygienist, denturist) | 3/468 | 1% |
|  | Dental assistant | 1/468 | 0.2% |
|  | Optometrist | 2/468 | 0.4% |
|  | Pharmacist or Registered Pharmacy Technician | 9/468 | 2% |
|  | Dietician/ Nutritionist | 5/468 | 1.1% |
|  | Natural healing practitioner (e.g. acupuncturist, traditional Chinese medicine (TCM) practitioner) | 7/468 | 1.5% |
|  | Other | 96/468 | 21% |
|  | No response | 20/468 | 4.3% |
| **Years of education/training** | 0-4 years | 218/468 | 47% |
|  | 5-9 years | 126/468 | 27% |
|  | 10+ years | 105/468 | 22.4% |
|  | Total respondents | 449/468 | 96% |
|  | No response | 19/468 | 4.1% |
| **Years of experience in most recent career** | 0-5 years | 63/468 | 13.5% |
|  | 6-10 years | 73/468 | 16% |
|  | 11-15 years | 78/468 | 17% |
|  | 16-20 years | 72/468 | 15.4% |
|  | 21-25 years | 64/468 | 14% |
|  | 26-30 years | 27/468 | 6% |
|  | 31-35 years | 44/468 | 9.4% |
|  | 36-39 years | 18/468 | 4% |
|  | 40+ years | 11/468 | 2.4% |
|  | Total respondents | 450/468 | 96.1% |
|  | No response | 18/468 | 4% |

About one-quarter of respondents (125/468, 27%) reported that their profession / area of occupation was nursing. Other commonly cited professions / areas of occupation were medical technologist / technician (32/468, 7%); nurse aide / orderly / patient services associate (28/468, 6%); occupational therapist (12/468, 3%); personal support worker (PSW) (12/468, 3%); paramedic (11/468, 2.4%) or allied primary health practitioner (10/468, 2.1%). About one-third of respondents (151/468, 32.3%) reported between 6 and 15 years of experience in their most recent career, close to one-third (136, 29.4%) between 16 and 25 years of experience, over one-fifth (100/468, 21.4%) over 26 years of experience, and a minority (63/468, 13.5%) 5 or fewer years. A large minority (218/468, 47%) reported 0-4 years of education / training, followed by close to one-third between 5-9 years (126/468, 27%), and over one-fifth (105/468, 22.4%) reporting 10 or more years of training. Small minorities reported not being in clinical practice (24/468, 5.1%) or holding an academic appointment / university affiliation (12/468, 3%). As to geographical location, most respondents (117/468, 25%) reported working in the Toronto health region, followed by the Southwest health region (121/468, 23%), Eastern region (85/468, 18.2%), Central region (76/468, 16.2%), and Northern region (40/468, 9%). Most participants (147/468, 31.5%) were employed full time, followed by those who reported being employed part time (97/468, 21%), self-employed (70/468, 15%), unemployed (70/468, 15%), casually employed (16/468, 3.4%) or a contractor (11/468, 2.4%) (Table 1).

### Vaccination Decision and Experiences

Most respondents (362/468, 77.4%) were not vaccinated (Chart 1). Of those who were vaccinated, most had completed a primary vaccine series (63/87, 72.4%), very few a partial primary series (13/87, 15%), and even fewer had been boosted once (5/63, 8%) or twice or more times (6/63, 9.5%). Most (65/87, 75%) were vaccinated primarily because it was mandated for work, a minority (14/87, 11%) to protect others, whether loved ones (6/87, 7%) or the larger community (4/87, 5%), and a smaller minority (4/87, 5%) to travel. Most vaccinated respondents (68/87, 78%) reported experiencing adverse effects post vaccination (National Cancer Institute, 2021). Adverse effects were *mild* after the 1^st^ dose (17/87, 20%), the 2^nd^ dose (17/87, 20%), or the 3^rd^ or more doses (3/87, 3.4%); *moderate* after 1^st^ dose (20/87, 23%), the 2^nd^ dose (8/87, 9.2%), or the 3^rd^ or more doses (3/87, 3.4%), or *serious* after the 1^st^ dose (8/87,9.2%), the 2^nd^ (15/87,17.2%), and the 3^rd^ or more doses (3/87, 3.4%). Close to one-third of vaccinated respondents (29/87, 33.3%) did not communicate their reaction to a doctor, while over one-third (31/87, 35.6%) did. Among these, in only a minority of cases (5/31, 16%) a report was filed, in most cases no report was filed (17/31, 55%), and in close to one-third of cases (9/31, 10%) respondents did not know if a report had been filed. Nearly one-third (27/87, 31%) of vaccinated respondents reported that after experiencing an adverse reaction their employer had still required additional doses. Only about one-fifth (19/87, 22%) reported no adverse events post vaccination ever (Table 2).

**Chart 1.**
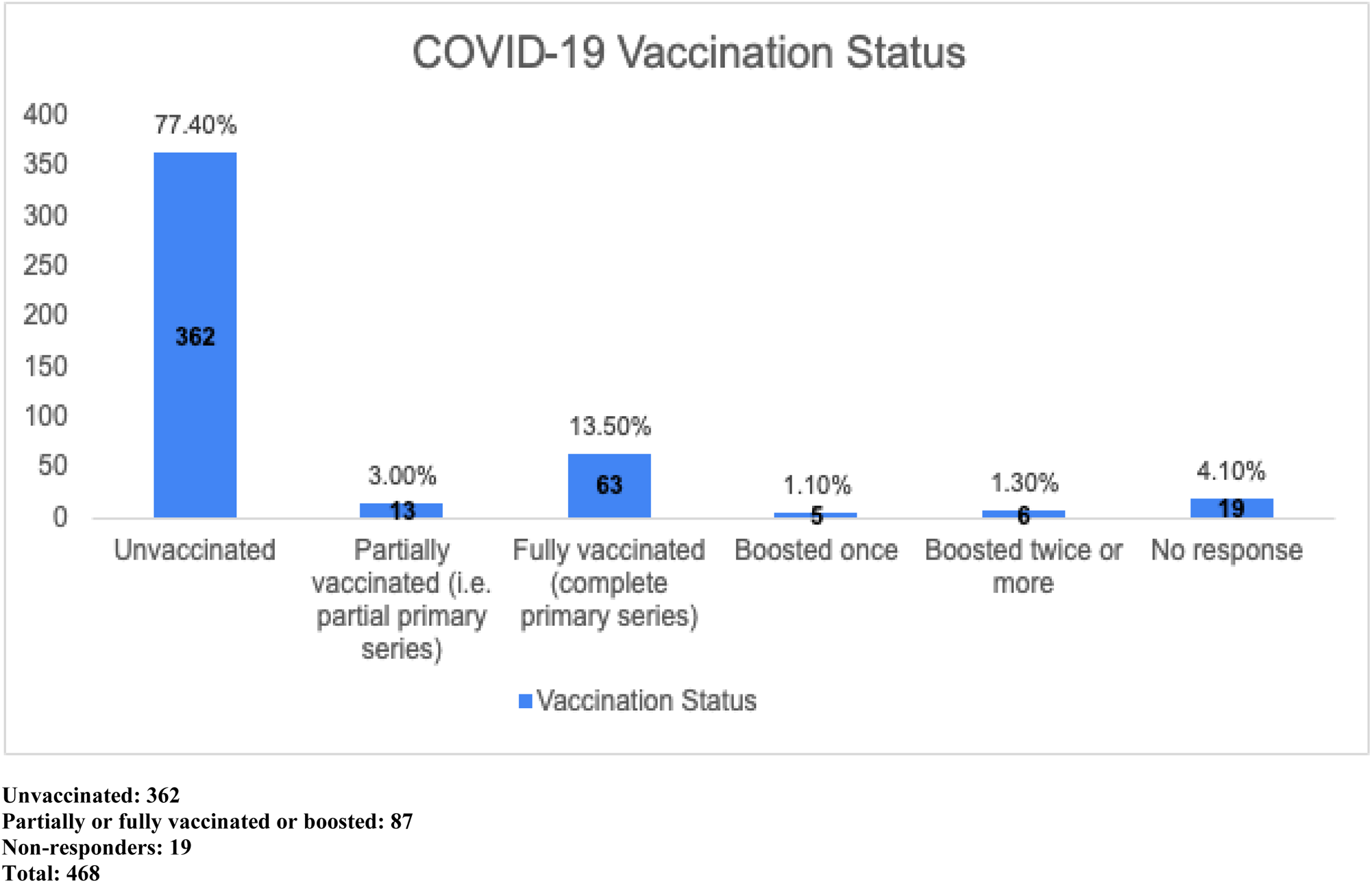
Vaccination status.

**Table 2.** Vaccination decision and experience^1^.

| **Question** | **Options** | **N** | **%** |
| --- | --- | --- | --- |
| Primary reason for getting vaccinated | To protect myself from severe outcomes (e.g., severe disease, hospitalization, death) | 1/87 | 1.1% |
|  | To protect my loved ones from severe outcomes (e.g., severe disease, hospitalization, death) | 6/87 | 7% |
|  | To protect the larger community from severe outcomes (e.g., severe disease, hospitalization, death) | 4/87 | 5% |
|  | It was mandated at work | 65/87 | 75% |
|  | It was mandated at school/ university | 1/87 | 1.1% |
|  | It was mandated at social venues (e.g., restaurants) | 0/87 | 0% |
|  | It was mandated for travel (e.g., visiting family or vacations) | 4/87 | 5% |
|  | It was mandated to visit vulnerable loved ones (e.g., grandparent in nursing home) | 1/87 | 1.1% |
|  | It was mandated at places of worship (e.g., church, temple, mosque) | 0/87 | 0% |
|  | I did it to avoid rejection from friends/ family members/ members of the community | 1/87 | 1.1% |
|  | Other | 3/87 | 3.4% |
|  | Total respondents | 86/87 | 99% |
|  | No response | 1/87 | 1.1% |
| Three main reasons for getting vaccinated | To protect myself from severe outcomes (e.g., severe disease, hospitalization, death) | 8/87 | 9.2% |

<sup>1</sup> Some answers allow for multiple options, so totals do not always amount to 100%

|  |  |  |  |
| --- | --- | --- | --- |
|  | To protect my loved ones from severe outcomes (e.g., severe disease, hospitalization, death) | 15/87 | 17.2% |
|  | To protect the larger community from severe outcomes (e.g., severe disease, hospitalization, death) | 14/87 | 16.1% |
|  | It was mandated at work | 75/87 | 86.2% |
|  | It was mandated at school/ university | 5/87 | 5.7% |
|  | It was mandated at social venues (e.g., restaurants) | 8/87 | 9.2% |
|  | It was mandated for travel (e.g., visiting family or vacations) | 23/87 | 26.4% |
|  | It was mandated to visit vulnerable loved ones (e.g., grandparent in nursing home) | 9/87 | 10.3% |
|  | It was mandated at places of worship (e.g., church, temple, mosque) | 1/87 | 1.1% |
|  | I did it to avoid rejection from friends/ family members/ members of the community | 7/87 | 8% |
|  | Other | 4/87 | 5% |
|  | No response | 0/87 | 81.3% |
| Vaccine side effects (if vaccinated) | N/A (no reactions ever) | 19/87 | 22% |
|  | Mild reaction after 1st dose | 17/87 | 20% |
|  | Mild reaction after 2nd dose | 17/87 | 20% |
|  | Mild reaction after 3rd or later dose | 3/87 | 3.4% |
|  | Moderate reaction after 1st dose | 20/87 | 23% |
|  | Moderate reaction after 2nd dose | 8/87 | 9.2% |
|  | Moderate reaction after 3rd or later dose | 3/87 | 3.4% |
|  | Severe reaction after 1st dose | 8/87 | 9.2% |
|  | Severe reaction after 2nd dose | 15/87 | 17.2% |
|  | Severe reaction after 3rd or later dose | 3/87 | 3.4% |
|  | Life-threatening after 1st dose | 1/87 | 1.1% |
|  | Life-threatening after 2nd dose | 0/87 | 0% |
|  | Life-threatening after 3rd dose or later dose | 0/87 | 0% |
|  | Other | 5/87 | 6% |
|  | No response | 0/87 | 82% |
| Communicated adverse reactions to GP / family doctor or other medical personnel | N/A (no adverse reaction) | 20/87 | 23% |
|  | No, I did not communicate my reaction to GP / other medical personnel | 29/87 | 33.3% |
|  | Yes, I communicated my reaction to GP / other medical personnel, and they filed a report | 5/87 | 6% |
|  | Yes, I communicated my reaction to GP / other medical personnel, but they did not file a report | 17/87 | 20% |
|  | Yes, I communicated my reaction to GP / other medical personnel, and I do not know if they filed a report | 9/87 | 10.3% |
|  | Other | 6/87 | 7% |
|  | Total respondents | 86/87 | 99% |
|  | No response | 1/87 | 1.1% |
| Experienced an adverse reaction after Covid-19 vaccine and still required to take additional doses | Yes | 27/87 | 31% |
|  | No | 48/87 | 55.2% |
|  | Prefer not to answer | 11/87 | 13% |
|  | Total respondents | 86/87 | 99% |
|  | No response | 1/87 | 1.1% |

### Personal and Family Impact of Vaccination Policies

Most respondents (311/468, 66.5%) agreed (50/468, 11%) or strongly agreed (261/468, 56%) that their current income was less than it was prior to the introduction of vaccination mandates. Not surprisingly, most laid off respondents (302/361, 84%) also agreed (47/361, 13%) or strongly agreed (257/361, 71.2%) that being terminated had significantly reduced their income, with about one-third of them (119/361, 33%) disagreeing (38/361, 11%) or strongly disagreeing (81/361, 22%) that they had needed to access financial supports or community services (e.g., food banks) upon losing their job. Responses to the impact of vaccination policies on physical health were more mixed. About half (224/468, 48%) of respondents chose “not applicable” - unsurprisingly since the majority of respondents were unvaccinated. A minority (78/468, 17%), however, reported that they agreed (24/468, 5.1%) or strongly agreed (56/468, 12%) that their physical health had worsened after mandates were implemented, while the remainder (144/468, 31%) were neutral (30/468, 6.4%), disagreed (60/468, 13%) or strongly disagreed (54/468, 12%) (Table 3).

**Table 3.**
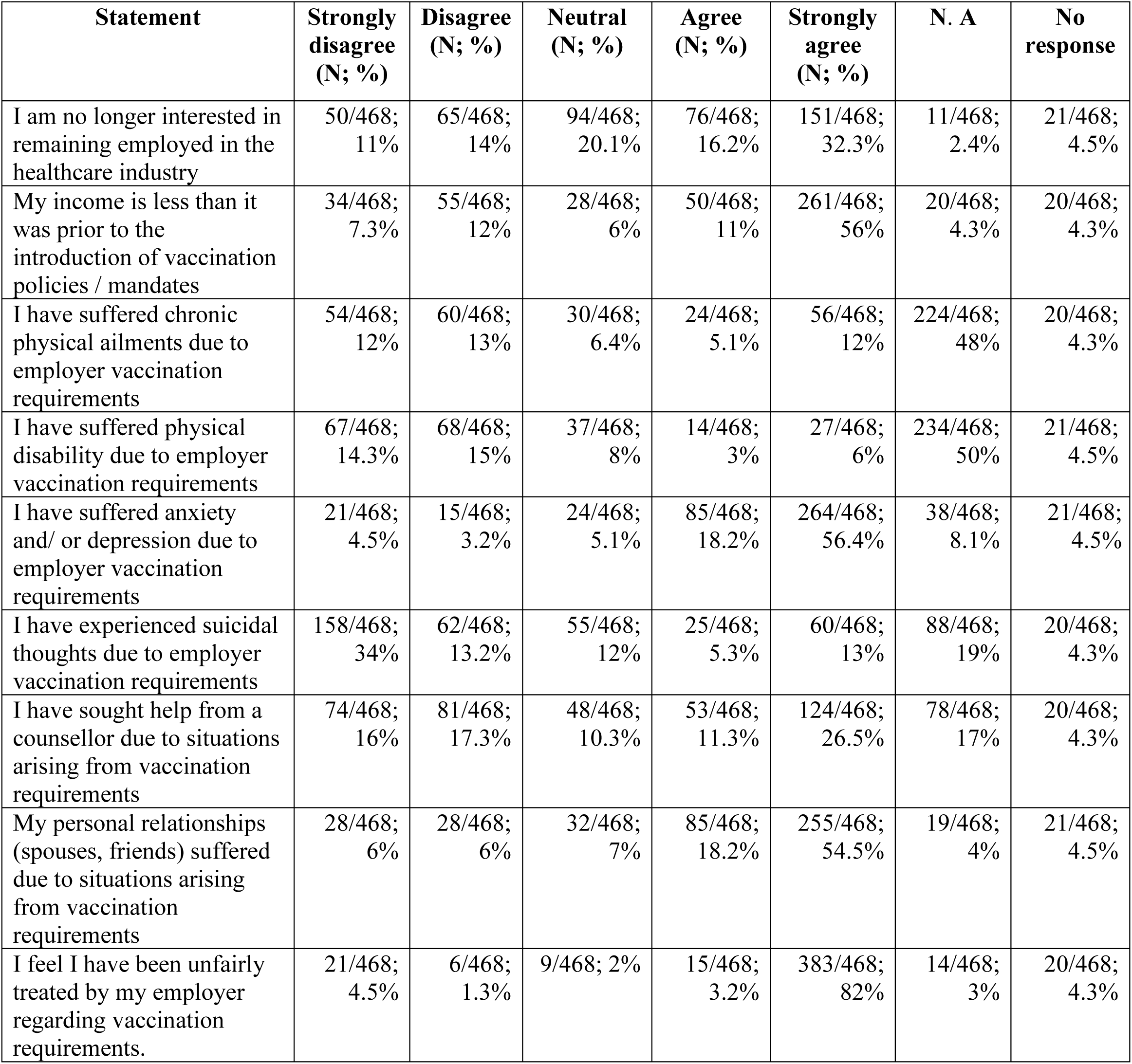
Personal impact of vaccination policies.

Similarly, answers to whether respondents had suffered physical disabilities due to employer vaccination requirements were also mixed, with close to one-third (135/468, 29%) disagreeing (68/468, 15%) or strongly disagreeing (67/468, 14.3%) that they had, about one-tenth (41/468, 9%) agreeing (14/468, 3%) or strongly agreeing (27/468, 6%), and a slightly smaller proportion (37/468, 8%) remaining neutral. In contrast, among respondents who reported that they had been terminated due to their vaccination decisions, most (219/361, 61%) either agreed (88/361, 24.4%) or strongly agreed (131/361, 36.3%) that losing their job had had a negative impact on their physical health. Even more pronounced was the impact of job termination on respondents’ mental health, with most (299/361, 83%) agreeing (97/361, 27%) or strongly agreeing (202/361, 56%) that being terminated had had a negative impact (Table 3).

Further, most respondents (349/468, 75%) reported experiencing anxiety or depression due to employer vaccination mandates, with close to one-fifth (85/468, 18.2%) agreeing (25/468, 5.3%) or strongly agreeing (60/468, 13%) that they had experienced suicidal thoughts due to employer vaccination requirements, and over one-third (177/468, 38%) agreeing (53/468, 11.3%) or strongly agreeing (124/468, 26.5%) that they had sought the help from a counsellor due to situations arising from these requirements. As well, most respondents (340/468, 73%) agreed (85/468, 18.2%) or strongly agreed (255/468, 54.5%) that their personal relationships had suffered due to situations arising from mandated vaccination, and most (398/468, 85%) agreed (15/468, 3.2%) or strongly agreed (383/468, 82%) with the statement “I feel I have been unfairly treated by my employer regarding vaccination requirements” (Table 3). Finally, while most respondents (280/468, 60%) reported good (127/468, 27.1%) or very good (153/468, 33%) physical health, over one-third (168/468, 36%) reported experiencing better physical health before Covid-19. This change was even more marked for mental health, that most (251/468, 54%) respondents rated as good (140/468, 30%) or very good (111/468, 24%), yet most (273/468, 58.3%) also rated their mental health as having been better before Covid-19 (Charts 2 & 3).

**Chart 2.**
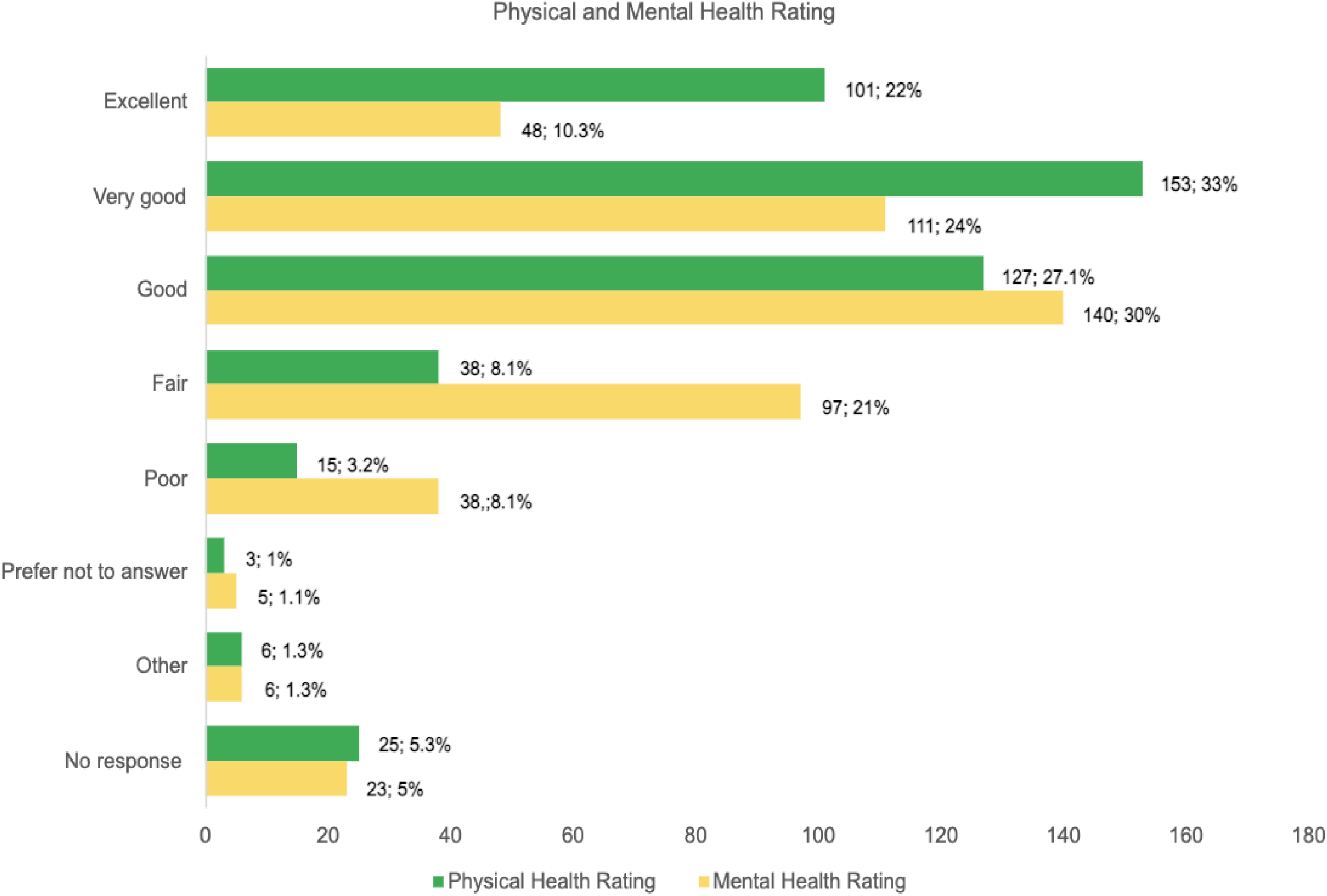
Physical and mental health self-rating.

**Chart 3.**
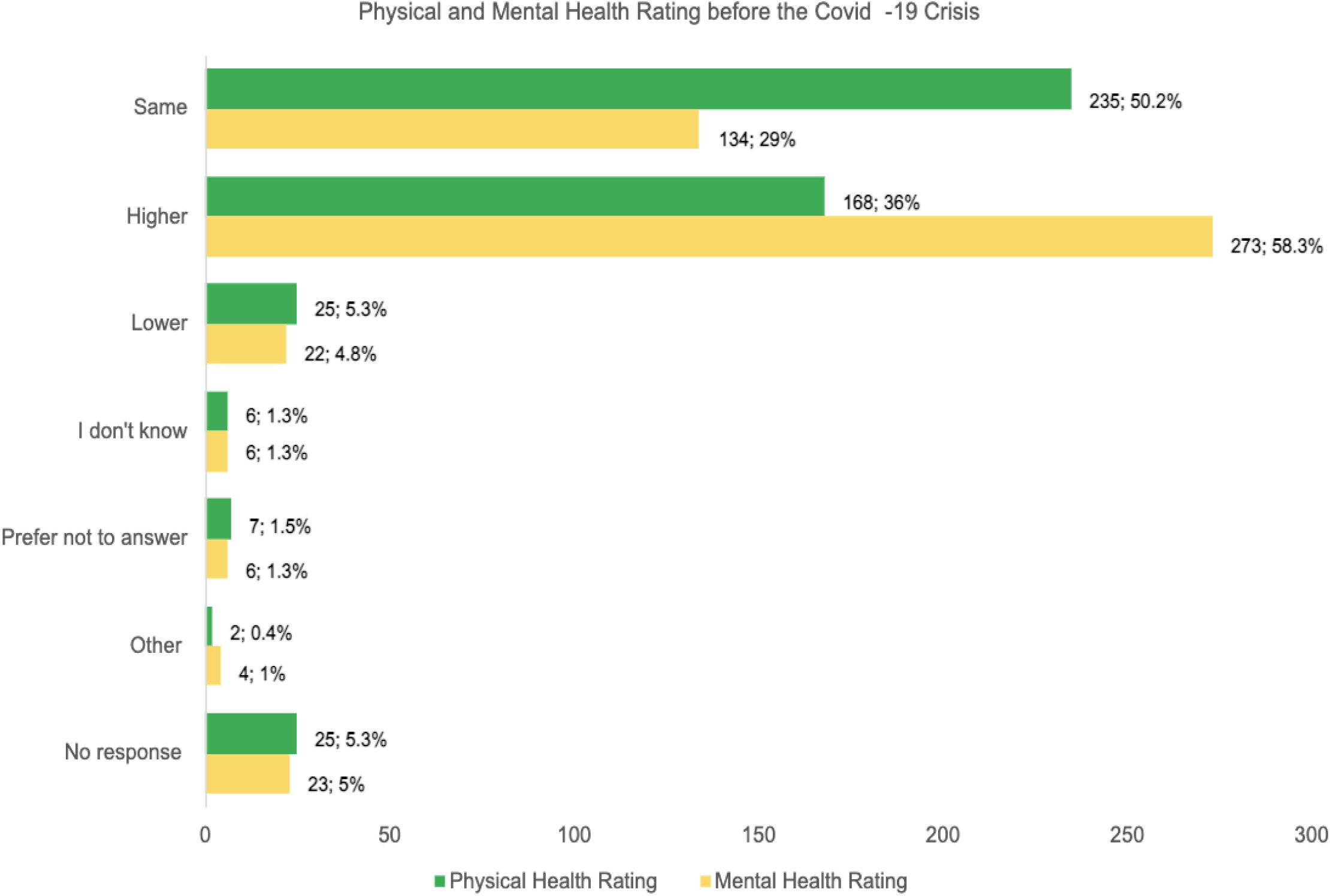
Physical and mental health self-rating before / after Covid-19. Interpretation: Over one third (N; 168; 36%) of respondents rated their physical health and more than half (N; 273; 58%) of respondents rated their mental health *higher* prior to Covid-19]

### Workplace and Labour Market Impact of Vaccination Policies

Nearly three-quarters (339/468, 72.4%) of respondents reported that they had been terminated due to their decision to not be vaccinated, either not at all or after one or two doses (i.e., booster mandates). In addition, about one-fifth (103/468, 22%) reported that they had been subjected to disciplinary measures other than termination, such as accusations of professional misconduct, reports to licensing colleges, temporary suspension of pay, exclusion from pension plans, or withdrawal of their professional license (Table 5). Finally, most respondents (351/468, 75%) agreed (80/468, 17.1%) or strongly agreed (271/468, 58%) that they had experienced conflict among colleagues, and between employees and management after the introduction of vaccines or vaccination policies. As well, most respondents (383/468, 82%) reported knowing of HCWs who had taken early retirement due to Covid-19 policies, most (397/468, 85%) knew of colleagues who had been laid off due to non-compliance with vaccine mandates, and most (385/468, 82.3%) knew of colleagues who had resigned because they did not wish to take the vaccine (Table 4).

**Table 4.** Vaccination requirements and impact on employment status and conditions.

| Question | Options | N | % |
| --- | --- | --- | --- |
| Terminated or laid off due to the decision to not receive the Covid-19 vaccine (first or subsequent doses)? | Yes | 339/468 | 72.4% |
|  | No | 88/468 | 19% |
|  | Prefer not to answer | 22/468 | 5% |
|  | Total respondents | 449/468 | 96% |
|  | No response | 19/468 | 4.1% |
| Subject to disciplinary measures *other* than layoffs (e.g., accusations of “professional misconduct”; reports to licensing colleges; temporary suspension of pay; exclusion from pension plan; withdrawal of professional license). | Yes | 103/468 | 22% |
|  | No | 274/468 | 56% |
|  | Prefer not to answer | 23/468 | 5% |
|  | Other | 49/468 | 10.5% |
|  | No response | 19/468 | 4.1% |
| Entitled to unemployment insurance if confirmed termination. | Yes | 27/361 | 7.5% |
|  | No | 315/361 | 87.3% |
|  | Prefer not to answer | 19/361 | 5.3% |
|  | Total respondents | 361/361 | 100% |
|  | No response | 0/361 | 0% |
| Offered alternatives in lieu of termination | Yes, but alternative was unaffordable (e.g., testing at my own expense) | 2/361 | 1% |
|  | Yes, but alternative was too cumbersome (e.g., testing every day) | 3/361 | 1% |
|  | No, I was not offered any alternative to vaccination | 342/361 | 95% |
|  | Other | 14/361 | 4% |
|  | Total respondents | 361/361 | 100% |
|  | No response | 0/361 | 0% |
| Found another job in their field | Yes | 145/361 | 40.2% |
|  | No | 195/361 | 54% |
|  | Prefer not to answer | 21/361 | 6% |
|  | Total respondents | 361/361 | 100% |
|  | No response | 0/361 | 0% |
| Retrained for a new career outside of the scope of healthcare | Yes | 95/361 | 26.3% |
|  | No | 248/361 | 69% |
|  | Prefer not to answer | 18/361 | 5% |
|  | Total respondents | 361/361 | 100% |
|  | No response | 0/361 | 0% |

**Table 5.**
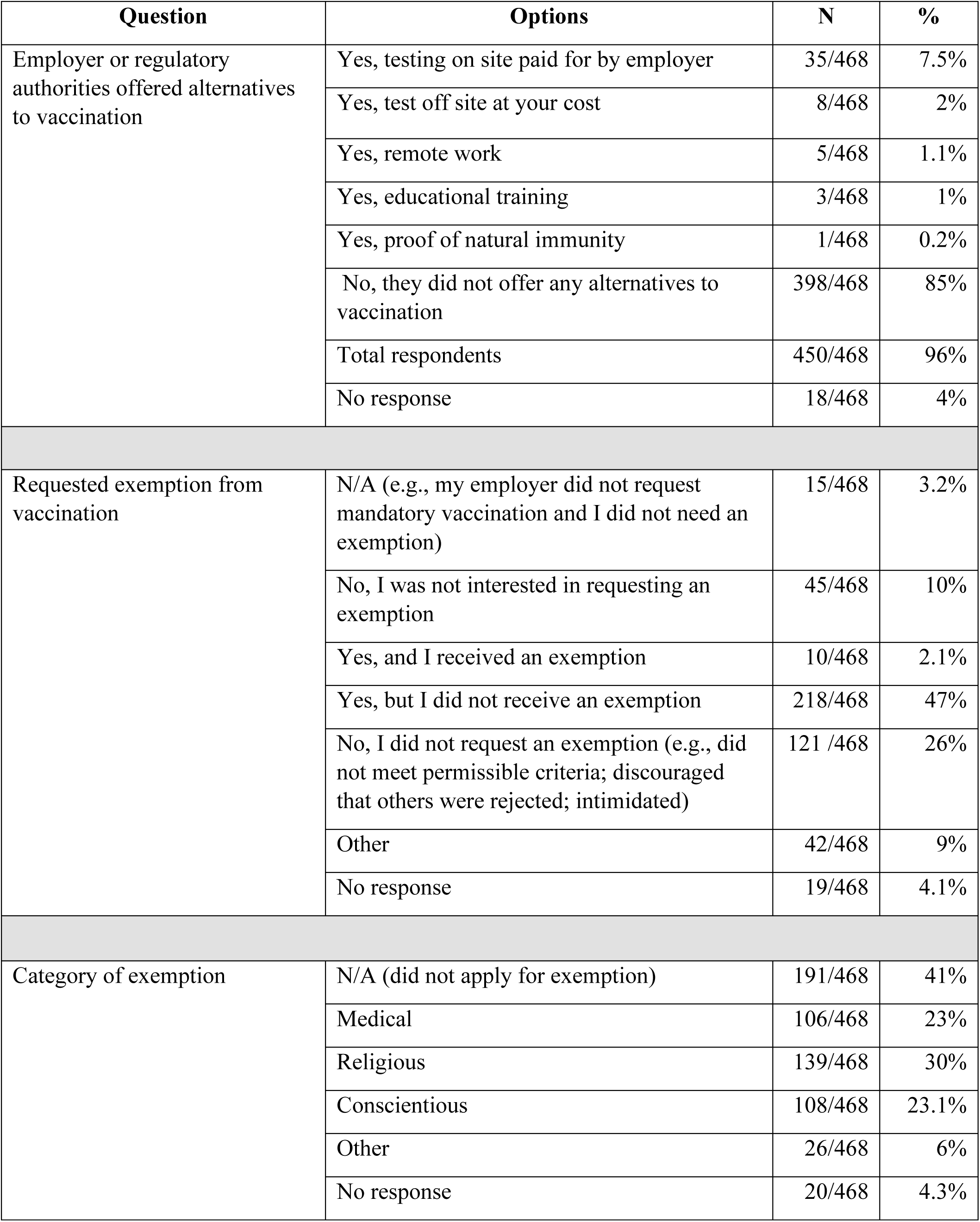
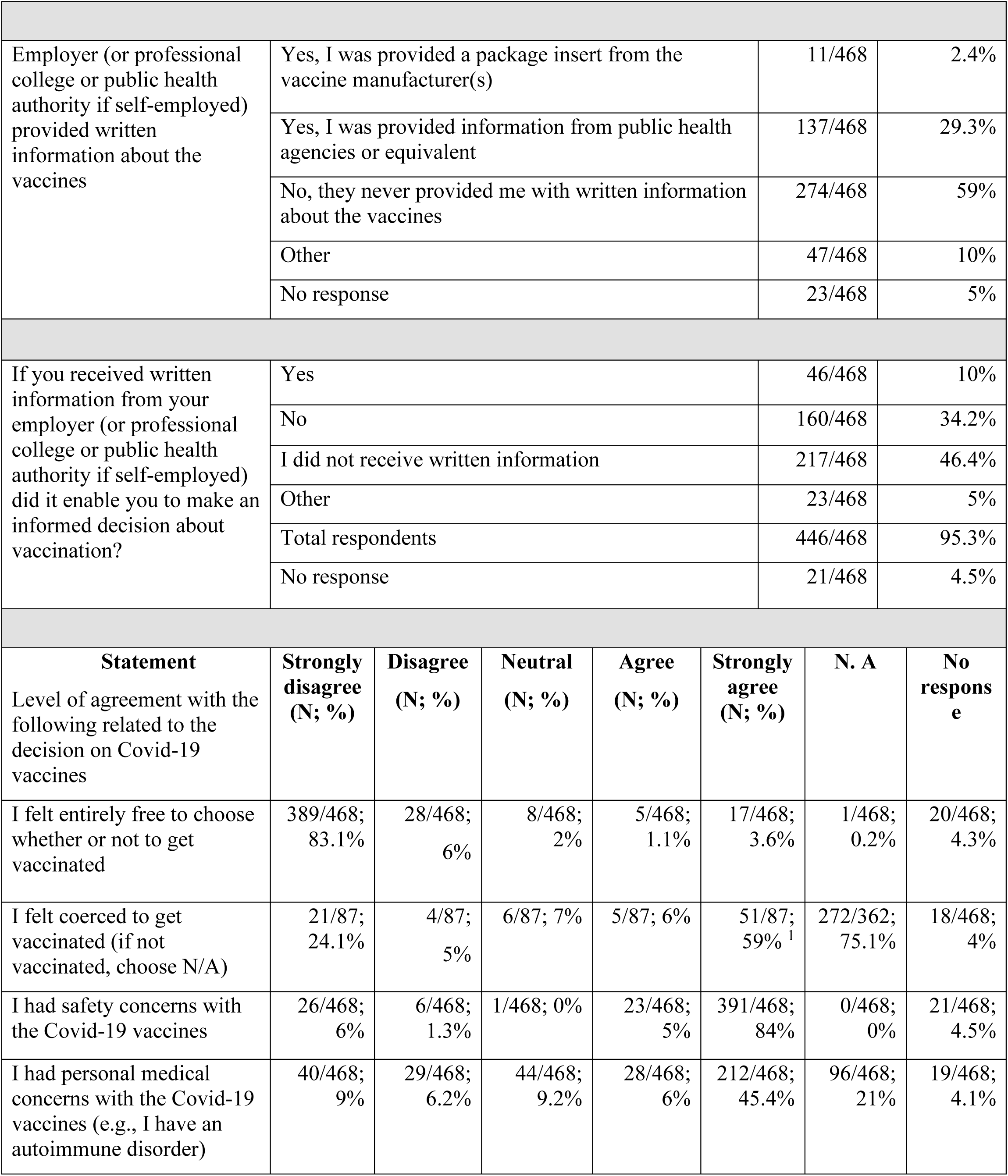
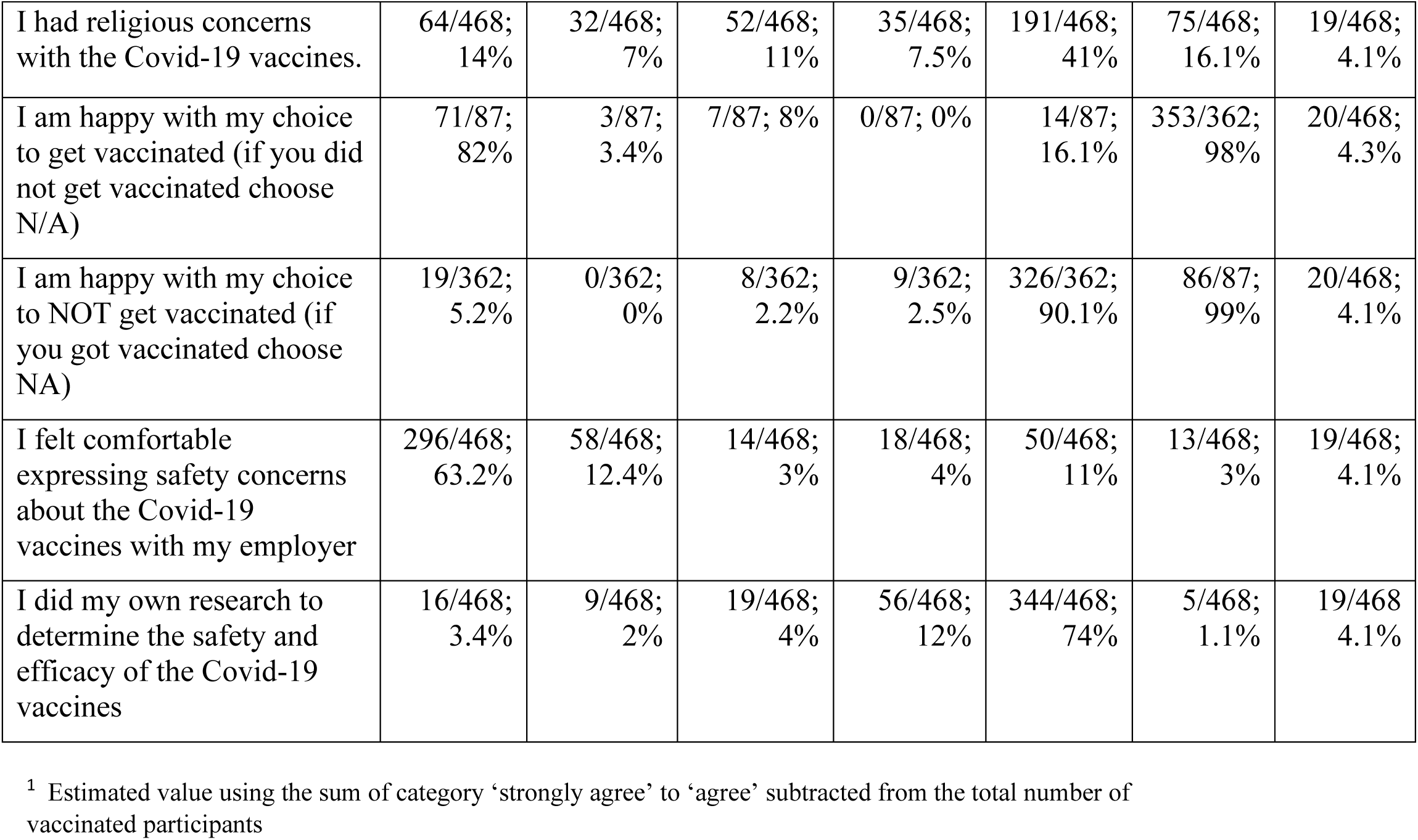
Accommodations, EDI considerations and informed consent.

In addition, about half (229/468, 49%) of respondents knew of students in the health professions who were de-enrolled by their educational institutions due to non-compliance with vaccination policies. The response to the statement “I would return to my previous role if possible / if mandates were dropped” was split. While over one-third (143/361, 40%) agreed (35/361, 10%) or strongly agreed (108/361, 30%) that they would, about the same proportion (134/361, 37.1%) disagreed (41/361, 11.4%) or strongly disagreed (93/361, 26%), while a small minority (59/361, 16.3%) was neutral. Finally, respondents reported mixed feelings about remaining employed in healthcare: close to half (199/468, 43%) agreed (58/468,12.4%) or strongly agreed (141/468, 30.1%) that they intended to leave their occupation or the healthcare sector altogether due to their experiences with Covid-19 policies, while a minority (76/468, 16.2%) reported they planned to remain in the industry, and about one-fifth (94/468, 20.1%) were neutral (Table 4).

### Accommodation, Equity Considerations & Informed Consent

Most respondents (398/468, 85%) reported that they were not offered any alternatives to vaccination. Nearly half (218/468, 47%) requested, but did not receive, an exemption. The most common reason for requesting an exemption was religious (139/468, 30%), followed by conscientious objection (108/468, 23.1%) and medical (106/468, 23%) grounds. However, over one-fourth of respondents (121/468, 26%) reported that they did not request an exemption because they were not eligible or felt intimidated or discouraged by the rejection of co-workers’ requests. When asked if their employer (or professional college or public health authority if self-employed) had provided them with written information about Covid-19 vaccines, most respondents (274/468, 59%) reported that they had not, almost one-third (137/468, 29.3%) reported that they were provided information from public health agencies or equivalent, and very few (11/468, 2.4%) reported being provided a package insert from the vaccine manufacturer. Over one-third (160/468, 34.2%) reported that, if received, the information from employers had not enabled them to make an informed decision about vaccination (Table 5).

When asked their level of agreement with a variety of statements related to informed consent, most respondents (417/468, 89.1%) disagreed (28/468, 6%) or strongly disagreed (389/468, 83.1%) that they had felt “entirely free to choose whether or not to get vaccinated”. In contrast, most (414/468, 88.5%) agreed (23/468, 5%) or strongly agreed (391/468, 84%) that they had safety concerns with Covid-19 vaccines, and more than half (260/468, 55.6%) agreed (28/468, 6%) or strongly agreed (212/468, 45.3%) that they had medical concerns, while close to half (226/468, 48.3%) agreed (35/468,7.5%) or strongly agreed (191/468, 41%) that they had religious concerns regarding the Covid-19 vaccines. Most respondents (354/468, 76%) also disagreed (58/468, 12.4%) or strongly disagreed (296/468, 63.2%) that they felt comfortable sharing their concerns with their employer, and most (400/468, 85.5%) agreed (56/468, 12%) or strongly agreed (344/468, 74%) that they did their own research regarding the safety and efficacy of Covid-19 vaccines. As well, most vaccinated respondents (56/87, 64.4%) agreed (5/87, 6%) or strongly agreed (51/87, 59%) that they had felt coerced to get vaccinated, and conversely, most (74/87, 85.1%) disagreed (3/87, 3.4%) or strongly disagreed (71/87, 82%) that they were happy with their choice to get vaccinated. In contrast, most unvaccinated respondents (335/362, 93%) agreed (9/362, 2.5%) or strongly agreed (326/362, 90.1%) that they were happy to have remained unvaccinated (Table 5).

### HCWs Views and Experiences of Mandates on Patient Care

Most respondents (351/468, 75%) had worked with Covid-19 positive or suspected patients prior to the vaccine mandate, and most (357/468, 76.3%) agreed (104/468, 22.2%) or strongly agreed (253/468, 54.1%) that they had observed concerning patient care or procedural changes upon the onset of Covid-19. Similarly, most (355/468, 76%) agreed (95/468, 20.3%) or strongly agreed (260/468, 56%) that they had observed disturbing patient care or procedural changes upon the introduction of Covid-19 vaccines, most (328/468,70.1%) agreed (69/468, 15%) or strongly agreed (259/468, 55.3%) that they had observed differential treatment of patients based on their vaccination status, and most (321/468, 69%) agreed (78/468, 17%) or strongly agreed (243/468, 52%) that they had observed an increase in patient harms associated with the Covid-19 vaccine. Significantly, only a very small minority (24/468, 5.1%) agreed (10/468/, 2.1%) or strongly agreed (14/468, 3%) that they had felt free to express to their employer their concerns about potential vaccine harms in patients, only a very small minority (36/468, 8%) agreed (11/468, 2.4%) or strongly agreed (25/468, 5.3%) that when they had expressed these concerns they had been documented or acted upon by their employer, and only a small minority (17/468, 4%) agreed (5/468,1.1%) or strongly agreed (12/468; 3%) that from the perspective of a potential patient, they felt confident that the healthcare system would provide adequate and quality care while respecting their personal preferences and values (Table 6).

**Table 6.** Impact on patient care.

| Question | Options |  |  |  |  | N | % |
| --- | --- | --- | --- | --- | --- | --- | --- |
| Worked with Covid-19 positive or suspected patients' pre-vaccine mandate | Yes |  |  |  |  | 351/468 | 75% |
|  | No |  |  |  |  | 51/468 | 11% |
|  | Not sure |  |  |  |  | 45/468 | 10% |
|  | Other |  |  |  |  | 3/468 | 1% |
|  | Total respondents |  |  |  |  | 450/468 | 96.2% |
|  | No response |  |  |  |  | 18/468 | 4% |
| Statement | Strongly disagree (N; %) | Disagree (N; %) | Neutral (N; %) | Agree (N; %) | Strongly agree (N; %) | N. A | No response |
| I observed concerning patient care or procedural changes coinciding with the onset of the Covid-19 crises | 10/468; 2.1% | 12/468; 3% | 35/468; 7.5% | 104/468; 22.2% | 253/468; 54.1% | 33/468; 7.1% | 21/468; 4.5% |
| I observed concerning patient care changes after the introduction of the Covid-19 vaccines | 7/468; 1.5% | 12/468; 3% | 27/468; 6% | 95/468; 20.3% | 260/468; 56% | 44/468; 9.4% | 23/468; 5% |
| I observed differential treatment of patients based on their vaccine status | 10/468; 2.1% | 14/468; 3% | 42/468; 9% | 69/468; 15% | 259/468; 55.3% | 53/468; 11.3% | 21/468; 4.5% |
| I observed an increase in patient harms associated with the Covid-19 vaccines | 7/468; 1.5% | 14/468; 3% | 49/468; 10.5% | 78/468; 17% | 243/468; 52% | 57/468; 12.2% | 20/468; 4.3% |
| I felt free to express any concerns I had about patient care or potential vaccine harms with my employer | 313/468; 67% | 52/468; 11.1% | 15/468; 3.2% | 10/468; 2.1% | 14/468; 3% | 45/468; 10% | 19/468; 4.1% |

| **Statement** | **Strongly disagree**<br>**(N; %)** | **Disagree**<br>**(N; %)** | **Neutral**<br>**(N; %)** | **Agree**<br>**(N; %)** | **Strongly agree (N; %)** | **N. A** | **No response** |
| --- | --- | --- | --- | --- | --- | --- | --- |
| From the perspective of a potential patient, I am confident that the current healthcare system will provide adequate and quality care while respecting my personal preferences and values | 342/468;<br>73.1% | 58/468;<br>12.4% | 24/468;<br>5.1% | 5/468;<br>1.1% | 12/468;<br>3% | 8/468;<br>2% | 19/468;<br>4.1% |
| I experienced conflict among colleagues at work after the introduction of vaccines and/or vaccination policies | 22/468;<br>5% | 27/468;<br>6% | 21/468;<br>4.5% | 78/468;<br>17% | 271/468;<br>58% | 28/468;<br>6% | 21/468;<br>4.5% |
| I experienced conflict between employees and management at work after the introduction of vaccines and/or vaccination policies | 21/468;<br>4.5% | 22/468;<br>5% | 25/268;<br>5.3% | 80/460;<br>17.1% | 270/468;<br>58% | 29/468;<br>6.2% | 21/468;<br>4.5% |
| I intend to leave or change my current job due to my experiences during the Covid-19 crises | 56/468;<br>12% | 47/468;<br>10% | 80/468;<br>17.1% | 43/468;<br>9.2% | 115/468;<br>25% | 108/468;<br>23.1% | 19/468;<br>4.1% |
| I intend to leave my occupation/ the healthcare sector / industry due to my experiences with the Covid-19 policy | 44/468;<br>9.4% | 60/468;<br>13% | 88/468;<br>19% | 58/468;<br>12.4% | 141/468;<br>30.1% | 56/468;<br>12% | 21/468;<br>4.5% |
| I was accused of undermining Covid-19 public health response / patient care due to my views / decisions about vaccination | 24/468;<br>5.1% | 35/468;<br>7.5% | 53/468;<br>11.3% | 81/468;<br>17.3% | 220/468;<br>47% | 36/468;<br>8% | 19/468;<br>4.1% |
| I was coerced into recommending / administering Covid-19 vaccines against my best clinical judgment (e.g., patient may experience an adverse event, or has experienced an adverse event post vaccination, Covid-19 or other; patient too young/old to benefit from vaccination, patient experienced Covid-19 and likely has strong natural immunity) | 44/468;<br>9.4% | 39/468;<br>8.3% | 43/468;<br>9.2% | 32/468;<br>7% | 102/468;<br>22% | 187/468;<br>40% | 21/468;<br>4.5% |
| I know of health workers who have taken early retirement due to Covid-19 policies | 22/468;<br>5% | 5/468;<br>1.1% | 14/468;<br>3% | 58/468;<br>12.4% | 325/468;<br>69.4% | 24/468;<br>5.1% | 20/468;<br>4.3% |
| I know of health workers who have been laid off due to failure to comply with vaccination | 19/468;<br>4.1% | 8/468;<br>2% | 10/468;<br>2.1% | 33/468;<br>7.1% | 364/468;<br>78% | 14/468;<br>3% | 20/468;<br>4.3% |
| I know of health workers who have resigned because they did not wish to take the vaccine | 21/468;<br>4.5% | 9/468;<br>2% | 13/468;<br>3% | 43/468;<br>9.2% | 342/468;<br>73.1% | 20/468;<br>4.3% | 20/468;<br>4.3% |
| I know of students in the health professions who were deregistered due to non-compliance with vaccination policies | 36/468;<br>9% | 36/468;<br>8% | 41/468;<br>9% | 23/468;<br>5% | 206/468;<br>44% | 106/468;<br>23% | 20/468;<br>4.3% |

| Question | Options | N | % |
| --- | --- | --- | --- |
| Encouraged to report adverse events post vaccination if observed | Yes | 21/468 | 4.5% |
|  | No | 423/468 | 90.4% |
|  | Total respondents | 444/468 | 95% |
|  | No response | 24/468 | 5.1% |
| Trained to report adverse events post-vaccination if observed | Yes | 20/468 | 4.3% |
|  | No | 424/468 | 91% |
|  | Total | 444/468 | 95% |
|  | No response | 24/468 | 5.1% |
| Administering Covid-19 vaccines part of job responsibilities | Yes | 31/468 | 7% |
|  | No | 412/468 | 88% |
|  | Total respondents | 443/468 | 95% |
|  | No response | 25/468 | 5.3% |
| Personally administered Covid-19 vaccines | Yes | 16/468 | 3.4% |
|  | No | 429/468 | 92% |
|  | Total respondents | 445/468 | 95.1% |
|  | No response | 23/468 | 5% |
| If administered Covid-19 vaccines, received compensation | Yes | 8/16 | 50% |
|  | No | 8/16 | 50% |
|  | Total respondents | 16/16 | 100% |
|  | No response | 0/16 | 0% |

| Statement (only HCWs who administered vaccines) | Strongly disagree (N; %) | Disagree (N; %) | Neutral (N; %) | Agree (N; %) | Strongly agree (N; %) | N. A | No response |
| --- | --- | --- | --- | --- | --- | --- | --- |
| I am aware that Covid-19 vaccines can cause serious or life-threatening injuries, including death | 0/16;<br>0% | 0/16;<br>0% | 1/16;<br>6.3% | 3/16;<br>19% | 12/16;<br>75% | 0/16;<br>0% | 0/16;<br>0% |
| I felt coerced to administer Covid-19 vaccines at any point during the vaccination campaign | 1/16;<br>6.3% | 0/16;<br>0% | 1/16;<br>6.3% | 1/16;<br>6.3% | 12/16;<br>75% | 1/16;<br>6.3% | 0/16;<br>0% |

As well, most (423/468, 90.1%) responded “no” when asked if they had been encouraged to report adverse events post vaccination if observed in patients. Nearly all (424/468, 91%) also reported “no” when asked if they had been trained to report adverse events post vaccination if observed. Significantly, close to one-third of respondents (134/468, 29%) agreed (32/468, 7%) or strongly agreed (102/468, 22%) that they had felt coerced to recommend / administer vaccines against their best clinical judgment. As well, most respondents (301/468, 64.3%) agreed (81/468, 17.3%) or strongly agreed (220/468, 47%) that they had been accused of undermining Covid-19 public health response / patient care due to their reservations about vaccination, and close to half (224/468, 4%) agreed (43/468, 9.2%) or strongly agreed (181/468, 39%) that they were disciplined for this reason. Finally, for a small number of respondents (16/468, 3.4%), job responsibilities included administering vaccines, and half of them (8/16, 50%) reported being reimbursed for the task (Table 9). Further, among the small number of respondents who had administered Covid-19 vaccines, most (15/16, 94%) agreed (3/16, 18.7%) or strongly agreed (12/16, 75%) that they believed that these can cause serious or life-threatening injuries, including death, and most (13/16, 81.3%) agreed (1/16, 6.2%) or strongly agreed (12/16, 75%) that they had felt coerced to administer Covid-19 vaccine despite their reservations (Table 6).

## Discussion

In our survey of 468 HCWs in the province of Ontario, Canada, most of them had 16 or more years of professional experience, were unvaccinated or failed to meet full vaccination requirements, and had been terminated due to non-compliance with mandates. As well, and regardless of vaccination status, most respondents reported safety concerns with vaccination, yet did not request an exemption due to their experience of high rejection rates by employers. However, most unvaccinated workers reported satisfaction with their vaccination choices, although they also reported significant, negative impacts of workplace mandates on their finances, their mental health, their social and personal relationships, and to a lesser degree, their physical health. In contrast, within the minority of vaccinated respondents, most reported being dissatisfied with their vaccination decisions, as well as having experienced mild to serious post vaccine adverse events, with about one-quarter within this group reporting having been coerced into taking further doses, under threat of termination, despite these events.

In relation to patient care, a large minority of respondents reported having witnessed underreporting or dismissal by hospital management of adverse events post vaccination among patients, worse treatment of unvaccinated patients, and concerning changes in practice protocols. Most respondents also reported being accused of undermining patient care due to their reservations about vaccination, and close to half reported being disciplined for this reason. Most respondents within the very small number those who actually administered Covid-19 vaccines reported that they had been coerced into doing so against their best clinical judgment. Finally, close to half of respondents reported their intention to leave the healthcare industry altogether.

This study has limitations. As with survey research generally, the sample size, while adequate as per usual practice standards (SurveyMonkey, n.d.), was nevertheless limited. As well, the types of respondents were limited by the ability of the research team and its professional connections to reach potential participants. For instance, most respondents were middle-income, women, and nursing professionals, only a handful were medical doctors, very few were low-paying - personal support workers, aides, or orderly – and only a small proportion self-identified as very low or very high income. It is likely that the policies impacted HCWs with different occupations and levels of income differentially. Our study is also cross-sectional, and we have only performed descriptive statistical analyses, so we are unable to observe the evolution of patterns across time, or relationships between variables, such as income and vaccination status. Very likely, there are income-level differences in HCWs’ ability to afford to not comply with the policy. There is, however, research suggesting that this may be the case. For example, a study of NHS elderly care home staff in England indicated that vaccine mandates led to lower rates of unvaccinated HCWs, especially among lower income workers (Girma & Paton, 2023) - unsurprisingly given that refusal was punished with job loss. That being said, better samples or more sophisticated methodological approaches would still be limited in their ability to address important legal and ethical challenges, that we elaborate on shortly.

There exists, to be sure, ample research on the perceived problem of “suboptimal” vaccine uptake or of vaccine hesitancy among HCWs, in Canada and elsewhere. As has been noted, most of this research, implicitly or explicitly, overwhelmingly endorses full and continuing vaccination for HCWs, if necessary through mandates (Chaufan et al., 2024). And there is little doubt that the policy of mandated vaccination has succeeded in increasing vaccination rates among HCWs (Lee et al., 2022; Poyiadji et al., 2022; Ritter et al., 2021). However, if the goal is to provide safer and better quality of care, then the success of the policy of mandated vaccination is questionable. For example, the study mentioned earlier, of NHS elderly care home staff in England, indicated *a reduction not only in rates of unvaccinated HCWs* but also of *between 3 and 4% in the healthcare labour force*, equivalent to 14,000 to 19,000 fewer HCWs, that had a negative impact on the health and well-being of residents in these establishments, thus undermining the ostensible goal of the policy (Girma & Paton, 2023).

Beyond pragmatic considerations such as impact on staff shortages, the policy of mandated vaccination for HCWs also has implications for law and ethics. Take the Canadian Constitution. Already in 1996, the *Canadian National Report on Immunization* outlined a lack of precedence for mandatory vaccination, noting that in contrast to other countries, “immunization is not mandatory in Canada; it cannot be made mandatory because of the Canadian Constitution” (Health Canada, 1997) (pg. 3). However, this provision of the report has been all but ignored in discussions around Covid-19 vaccination mandates. So apparently has been other important Canadian legislation, for instance, the 1996 *Health Care Consent Act,* which states that medical treatments should not be administered without the voluntary consent of individual recipients (Health Care Consent Act, 1996, S.O. 1996, c. 2, Sched. A, 2014), and the 2004 *Personal Health Information Protection Act*, which states that consent for disclosure of personal information cannot be obtained through coercion (Personal Health Information Protection Act, 2004, S.O. 2004, c. 3, Sched. A, 2014).

Nevertheless, in recent years at least some courts have sided in favour of terminated HCWs in Ontario. One arbitrator, for instance, noted that the respondent, a medical establishment, had failed to consider evidence for the inability of Covid-19 vaccines to prevent transmission, and to grant accommodations for unvaccinated employees. One of the grounds for the arbitrator’s decisions was that hospital management, given mounting evidence for waning vaccine effectiveness, had decided against a booster mandate because the alleged benefits of boosters did not outweigh the risk of further staff losses. However, noted the arbitrator, current policy had still permitted the hospital to retain employees with only two vaccine doses, *despite management’s acknowledgment of their waning effectiveness*, while unvaccinated workers were terminated. An additional ground for siding in favour of terminated HCWs was that one of the plaintiffs had been fired after returning from parental leave in April 2023, long after evidence of waning effectiveness was available and admitted by hospital managers themselves. Also referenced in the legal proceedings were hospital statistics recording that most Covid-19 infections had occurred among fully vaccinated staff, thus the conclusion of the arbitrator that the policy was “unreasonable” (Quinte Health v Ontario Nurses Association, 2024).

In another case pertaining to two HCWs in clerical positions, the arbitrator also sided with the HCWs upon concluding that, in their refusing to reveal their vaccination status, they were “exercising their right to choose whether to receive medical treatment and […] whether to disclose private medical information” […], that even a “reasonable vaccination policy […] does not eliminate the grievors’ right of medical consent”, [and that] the mere fact that the [HCWs] were unwilling to have a vaccine injected into their bodies cannot fairly be characterized as an act of insubordination, or some other culpable conduct”, and, for this reason, termination was “not a permissible employer response to the exercise of this right” (Humber River Hospital v Teamsters Local Union No. 419, 2024) (pg. 4).

Similar legal issues have been debated elsewhere, specifically on the matter of the weak or non-existent liability of vaccine manufacturers generally. For example, Holland has described models for vaccine injury liability in the USA and European Union, detailing how, due to existing legislation and judicial decisions, vaccine makers in the USA have “almost blanket liability protection from damages for vaccine harms”, in contrast to the European Union, where, based on a ruling from the European Court of Justice (ECJ), someone injured by a vaccine has the right to compensation via a civil court, even if “a scientific consensus that a vaccine can cause the alleged injury does not yet exist” (Holland, 2018) (pg. 416). The author proposed that making vaccine manufacturers legally liable for potential harms caused by their products would “better balance the concerns of public health and individual rights”, and therefore strengthen both vaccine safety and confidence (pg. 417). The same could be said about Canada, where, since 2017, all vaccine manufacturers, including Covid-19 vaccine manufacturers, have been legally immune to charges against them stemming from any harm caused by their products (Gilmore, 2020).

Nevertheless, there is little doubts that these harms exist, and evidence for them continues to accumulate (Buchan et al., 2022; Fraiman et al., 2022; Karlstad et al., 2022; Li et al., 2021; Mansanguan et al., 2022). Identified adverse events post vaccination, such as myopericarditis upon receiving mRNA vaccines (Naveed et al., 2022; Public Health Agency of Canada, 2021) and lack of reproductive toxicity data, admitted even by national public health authorities (UK.Gov, 2022), are especially relevant to young populations. More recently, one study of over 99 million participants indicated an observed vs. expected (OE) ratio for acute disseminated encephalomyelitis of 3.78, for cerebral venous sinus thrombosis of 3.23, and for Guillain-Barré syndrome of 2.49, and an excess risk of serious adverse events of special interest, including death, between 10.1 and 15.1 (Faksova et al., 2024).

As well, a recent report by the *National Academies of Sciences, Engineering, and Medicine* (NASEM), upon prefacing that the benefits of (all) vaccines are “well-established”, reported “convincing evidence” of a causal relationship between mRNA Covid-19 vaccines and myocarditis, “inadequate [evidence] to accept or reject a causal relationship between mRNA-1273 (Moderna) and ischemic stroke”, evidence for accepting “a causal relationship between Astra Zeneca Covid-19 vaccines” and two specific adverse effects, thrombosis with thrombocytopenia syndrome and Guillain-Barré syndrome, and evidence for multiple vaccines and various shoulder injuries (National Academies of Sciences, Engineering, and Medicine, 2024) (pg. 5). Finally, a report from the World Council for Health based on data from several reporting systems - VigiAccess Database (WHO), the Vaccine Adverse Event Reporting System (USA; CDC, FDA), Eudravigilance (European Medicines Agency), and the UK Yellow Card Scheme (NHS) - found an “unprecedented” number of reports of adverse events in all the included databases, along with evidence for waning, or lack of, effectiveness, concluding that Covid-19 vaccines should be recalled (World Council for Health, 2022). Our research provides support for these scientific, legal, and ethical challenges.

## Conclusions

In 2021 the Organization for Economic Cooperation and Development (OECD) announced six evaluation criteria that jointly provide “a normative framework […] to determine the merit or worth of an intervention” - a policy, a strategy, or an activity (OECD, 2021). The first criterion is “relevance”, i.e., to what extent a policy is responsive to beneficiaries, meaning those who “benefit directly or indirectly from the policy”. The second criterion is “coherence”, i.e., to what extent a policy is compatible with other policies in a given setting. The third is “effectiveness”, i.e., to what extent a policy has achieved or is expected to achieve its objectives”. The fourth criterion is “efficiency”, to what extent a policy converts inputs into outputs in the “most cost-effective way possible, as compared to feasible alternatives in the context” and within a reasonable timeframe. The fifth criterion is “impact”, i.e., to what extent a policy “has generated or is expected to generate significant positive or negative, intended or unintended”, effects. The sixth and last criterion is “sustainability”, i.e., whether benefits are likely to last (OECD, 2021).

If our findings indicate a trend in the health care sector in Ontario, Canada, they suggest that by these criteria the policy of mandated vaccination for HCWs in the province has failed in its purported goal of promoting safer healthcare environments and achieving better care. Concerning “relevance”, the intended beneficiaries, whether HCWs, patients, or communities at large, have been harmed by exacerbated staff shortages, intimidating work environments, and health professionals coerced into acting against their best clinical judgment. Concerning “coherence”, the policy has proven to be at odds with other policies within health settings, such as the imperative to maintain adequate staffing levels or to respect informed consent and bodily autonomy, not only for HCWs but for patients who, for whatever reason, decline vaccination. As to “effectiveness”, there is no evidence that the policy has improved patient care – as suggested by our findings, it has likely worsened it.

Concerning “efficiency”, there is no evidence that the policy has been more cost-effective than comparable alternatives, such as relying on the superiority of natural immunity over artificial immunity (Chemaitelly et al., 2021, 2022; Gazit et al., 2022; Hall et al., 2021), acquired by most HCWs during 2020 as they treated patients in critical need, and for this reason were celebrated as heroes by the media and the authorities (Office of the Premier, 2020; Simmons, 2020). In fact, there is no evidence that such (then unvaccinated) workers were deemed a threat to patient safety and disciplined for that reason. Concerning “impact”, our findings also suggest that the overall impact of the policy on the well-being of HCWs and the sustainability of health systems has also been negative. Finally, concerning “sustainability”, with close to half of our sample of highly trained and experienced HCWs intending to leave the health professions, we see no evidence for any net benefits, either current or future. We conclude that if, by the OECD criteria, the policy of mandated vaccination for HCWs has failed, this failure, along with the contested efficacy and safety of Covid-19 vaccines, their negative impact on HCWs’ wellbeing, staffing levels, and patient care, and the threat that mandates represent to longstanding bioethical principles such as informed consent and bodily autonomy (UNESCO, 2005; World Medical Association, 1964), negates any basis - policy, scientific, or ethical - to continue with the practice.

## Data Availability

All data produced in the present work are contained in the manuscript

## Author contributions

C.CH designed the project, oversaw the data collection and analysis, and drafted the first version of the manuscript. NH conducted the literature review and made substantive intellectual contributions. NH and RM assisted with data collection and analysis and contributed to subsequent versions of the manuscript. All authors have read and approved the current version.

## Acknowledgments

CC thanks the many professional and lay organizations, students, trainees, and friends who have afforded spaces of reflection and debate over the past years, and especially her husband Julian Field, for his editorial feedback and support. NH thanks her family and friends for their ongoing encouragement and support, and Dr. Chaufan for her mentorship. RM thanks her friends, family, and Dr. Chaufan for their unwavering support and guidance. All authors are grateful to the participants for sharing with us their life experiences and making this study possible.

## Funding

This work was funded by a New Frontiers in Research Fund (NFRF) 2022 Special Call, NFRFR-2022-00305.

## Footnotes

1 Percentages in the tables were rounded to the first decimal point, which may explain minor inconsistencies among categories.

## Notes

### Competing Interest Statement

The authors have declared no competing interest.

### Author Declarations

Institutional Review Board, York University, Canada

